# Risk factors associated with mortality of COVID-19 in 3125 counties of the United States

**DOI:** 10.1101/2020.05.18.20105544

**Authors:** Ting Tian, Jingwen Zhang, Liyuan Hu, Yukang Jiang, Congyuan Duan, Zhongfei Li, Xueqin Wang, Heping Zhang

## Abstract

**Background:** The number of cumulative confirmed cases of COVID-19 in the United States has risen sharply since March 2020. A county health ranking and roadmaps program has been established to identify factors associated with disparity in mobility and mortality of COVID-19 in all counties in the United States.

**Methods:** To find out the risk factors associated with county-level mortality of COVID-19 with various levels of prevalence, a negative binomial design was applied to the county-level mortality counts of COVID-19 as of August 27, 2020 in the United States. In this design, the infected counties were categorized into three levels of infections using clustering analysis based on time-varying cumulative confirmed cases from March 1 to August 27, 2020. COVID-19 patients were not analyzed individually but were aggregated at the county-level, where the county-level deaths of COVID-19 confirmed by the local health agencies.

**Results:** 3125 infected counties were assigned into three classes corresponding to low, median, and high prevalence levels of infection. Several risk factors were significantly associated with the mortality counts of COVID-19, where higher level of air pollution (0.153, *P*<0.001) increased the mortality in the low prevalence counties and elder individuals were more vulnerable in both the median and high prevalence counties. The segregation between non-Whites and Whites and higher Hispanic population had higher likelihood of risk of the deaths in all infected counties.

**Conclusions:** The mortality of COVID-19 depended on sex, race/ethnicity, and outdoor environment. The increasing awareness of the impact of these significant factors may lead to the reduction in the mortality of COVID-19.

## Introduction

COVID-19 is an infectious disease caused by a novel coronavirus with an estimated average incubation period of 5.1 days[1]. It spreads through person-to-person transmission, and has now infected 215 countries and regions with over 24 million total confirmed cases as of August 27, 2020[2]. The United States had 5,867,785 confirmed cases on August 27, 2020, the highest in the world, but there were only 69 confirmed cases on March 1, 2020[3].

The United States has been suffering from a severe epidemic, with COVID-19 related deaths occurring all over the country. For instance, New York City had the largest number of total deaths (23,674), accounting for the majority of deaths in the infected counties, while no one in King county, Texas was infected as of August 27, 2020[3]. Therefore, it is of great interest to find out the risk factors that influence the number of deaths of COVID-19. It is known that infectious diseases are affected by factors other than medical treatments[4, 5]. For example, influenza A is associated with obesity[6], and the spread of the 2003 SARS events depends on seasonal temperature changes[7].

The County Health Rankings and Roadmaps program was launched by both the Robert Wood Johnson Foundation and the University of Wisconsin Population Health Institute[8]. This program has been providing annual sustainable source data including health outcomes, health behaviors, clinical care, social and economic factors, physical environment and demographics since 2010. We explored putative risk factors that may affect the mortality of COVID-19 in different areas of the United States in order to increase awareness of the disparity and aid the development of risk reduction strategies.

## Methods

### Data sources

We collected the number of cumulative confirmed cases and deaths from March 1 to August 27, 2020, for counties in the United States from the New York Times[9]. The COVID-19 confirmed cases and deaths were identified by the laboratory RNA test and specific criteria for symptoms and exposures from health departments and U.S. Centers for Disease Control and Prevention (CDC). The county health rankings reports from year 2020 were compiled from the County Health Rankings and Roadmaps program official website[8]. There were 77 measures in each of 3142 counties, including the health outcome, health behaviors, clinical care, social and economic factors, physical environment, and demographics. We refer to the official website of the County Health Rankings and Roadmaps program[8] for detailed information.

### Study areas

As of August 27, 2020, a total of 3,208 counties reported confirmed cases in the United States, leaving 3125 counties with both confirmed cases of COVID-19 and county health ranking data recorded to be analyzed in this study. The total number of deaths as of August 27, 2020 was considered as the outcome of this study.

### Assessment of covariates in health factors

We divided the putative risk factors[8] into 5 categories: health behaviors (e.g., access to exercise opportunities, insufficient sleep), clinical care (e.g. primary care physicians ratio), social and economic factors (e.g., racial segregation index), physical environment (e.g., transit problems and air quality), and demographics (age, sex, rural, and race/ethnicity). For example, there were previous studies which identified the air pollution may relate to high levels of COVID-19[10] and elder population had the high risk in the COVID-19[11]. Besides these identified risk factors, we were interested in the adverse health factors may link to the mortality of COVID-19. Table 1 presented descriptive definition, sources and literature of 12 risk factors. All deaths resulted from complications of COVID-19.

**Table 1.**
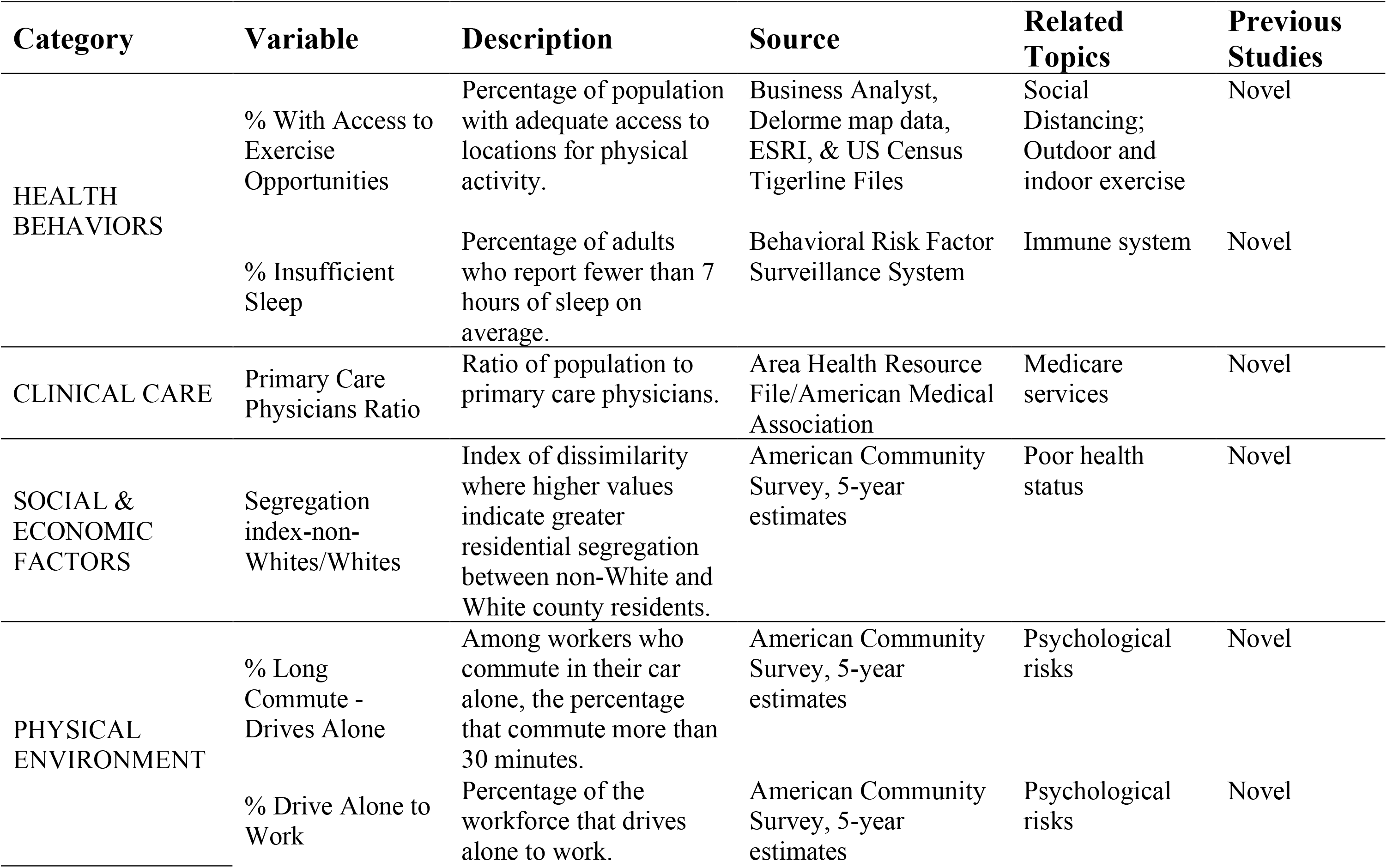
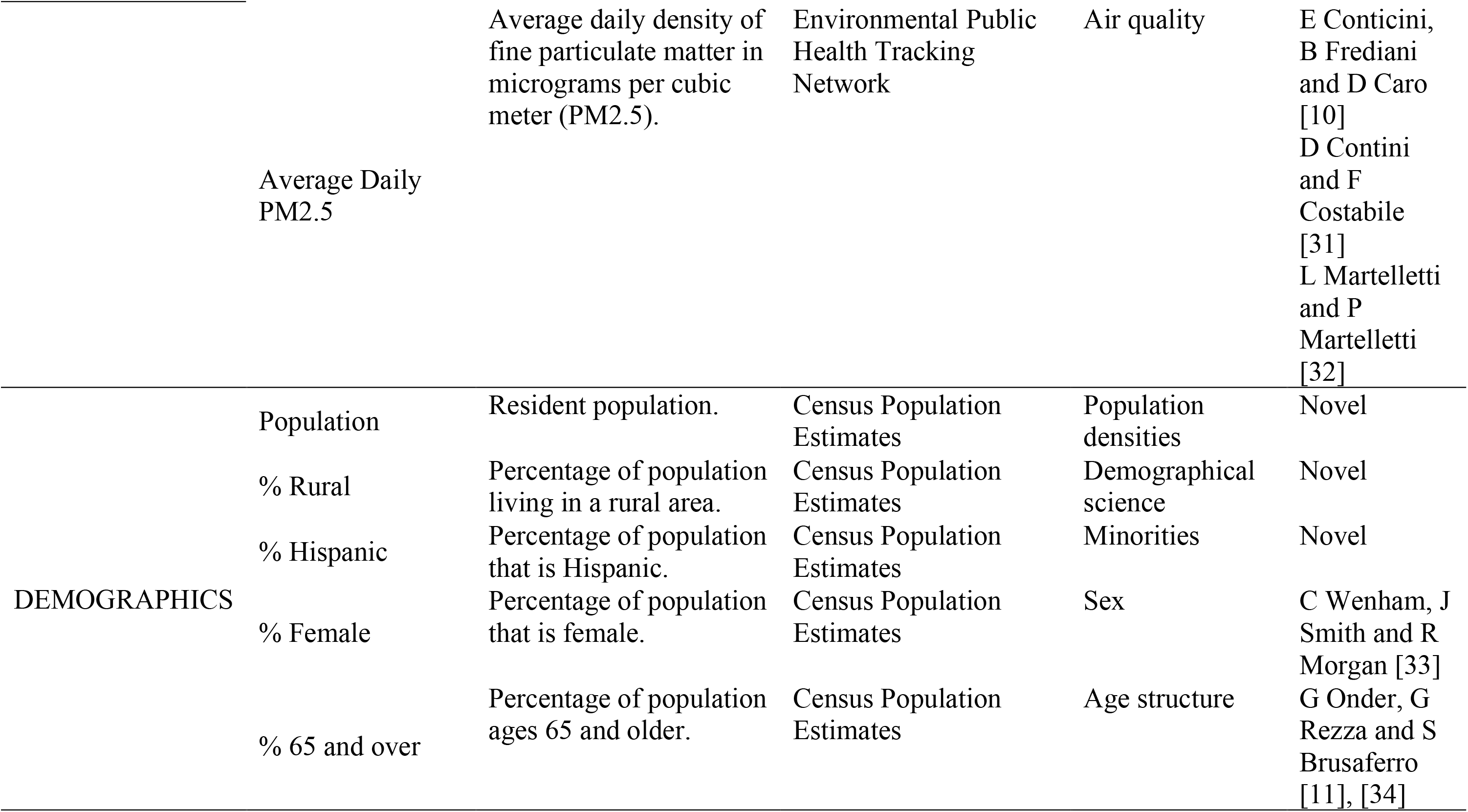
The definitions and sources of the 12 selected risk factors, and the literature supporting these factors.

### Statistical analysis

The trend of the cumulative confirmed cases varied greatly in counties of the United States. We used the partitioning around medoids (PAM) clustering algorithm[12, 13] to assign counties with similar trends into a homogenous class after standardizing the time series of cumulative confirmed cases from March 1 to August 27, 2020. Based on the clustering results, we used the Kruskal-Wallis test[14] to detect whether there were significant differences in the distributions of 12 risk factors across different classes of counties. The 12 risk factors were used to build a negative binomial model[15, 16] for every class of the counties. The analysis was conducted in R version 3.6.1.

### Validation analysis

We randomly divided counties (samples) into training (70% of the counties) and testing (30% of the counties) in each class. The model obtained from the training data was employed to predict the death counts of COVID-19 in the testing data, and the accuracy was assessed by the root mean square error (RMSE) of the mortality ratio (the number of deaths divided by the number of cumulative confirmed cases).

## Results

### Three classes of county-level infection in the United States

The clustering analysis grouped the 3,125 counties were assigned into 3 classes. There were 2,751 counties in the first class with the lowest overall cumulative confirmed cases. Its medoid was Halifax County in Virginia. There were 294 counties in the second class with a median level of overall cumulative confirmed cases. Its medoid was St. Clair County in Illinois. There were 80 counties in the third class with the highest overall cumulative confirmed cases. Its medoid was Marion County in Indiana. Here, the PAM algorithm selected the county with most representative data as the medoid in a class[12, 13]. The geographical distribution of the counties by class was shown in Figure 1, where the size of a circle indicated the cumulative confirmed cases on August 27, 2020. The distribution of deaths on August 27, 2020, which clearly differed among the three classes, was also presented in Figure 1. Note that the east, south, and west coasts were the most severely hit areas by COVID-19. Most counties in the high prevalence class were from Massachusetts, New York, New Jersey, Florida, Texas and California [9].

**Figure 1.**
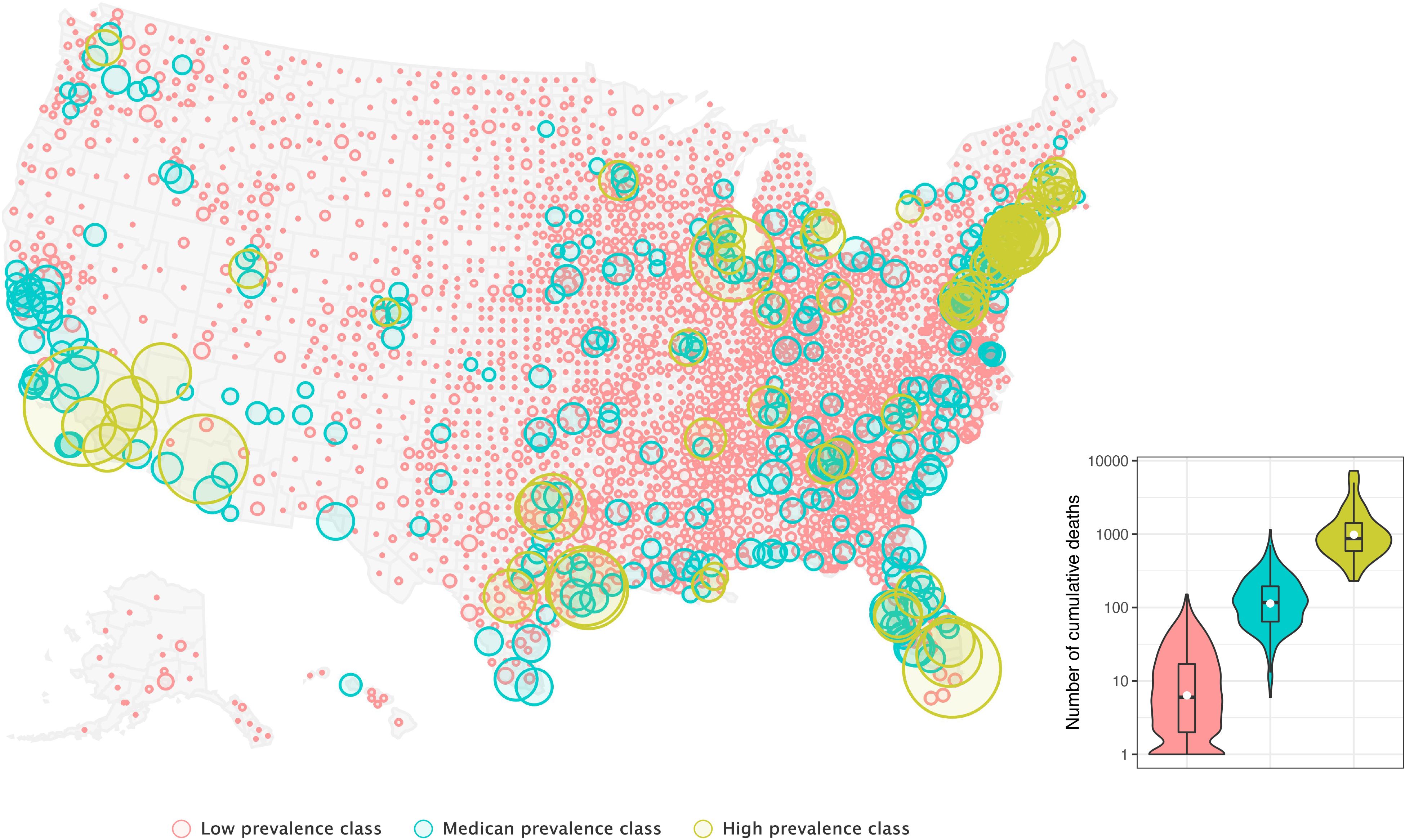
The geographical distribution of three classes of counties. The clustering was based on time-variant cumulative confirmed cases from March 1 to August 27, 2020. The size of circle represented the total confirmed cases on August 27, 2020. The distributions of deaths on August 27, 2020 in the three classes of counties were combined.

### Distributions of 12 selected risk factors in the three classes of counties

Figure 2 showed the distributions of the 12 selected risk factors by the class of counties. The distributions were significant different (*P*<0.001) for all 12 risk factors. For example, the average population in the low prevalence class was 38,444, which was 10% and 3% of the average populations in the median and high prevalence classes, respectively. The average proportion of rural residents in the low prevalence class was 64.47%, versus 2.72% in the high prevalence class. The segregation index of non-Whites versus Whites was the largest in the high prevalence class, but the smallest in the low prevalence class.

**Figure 2.**
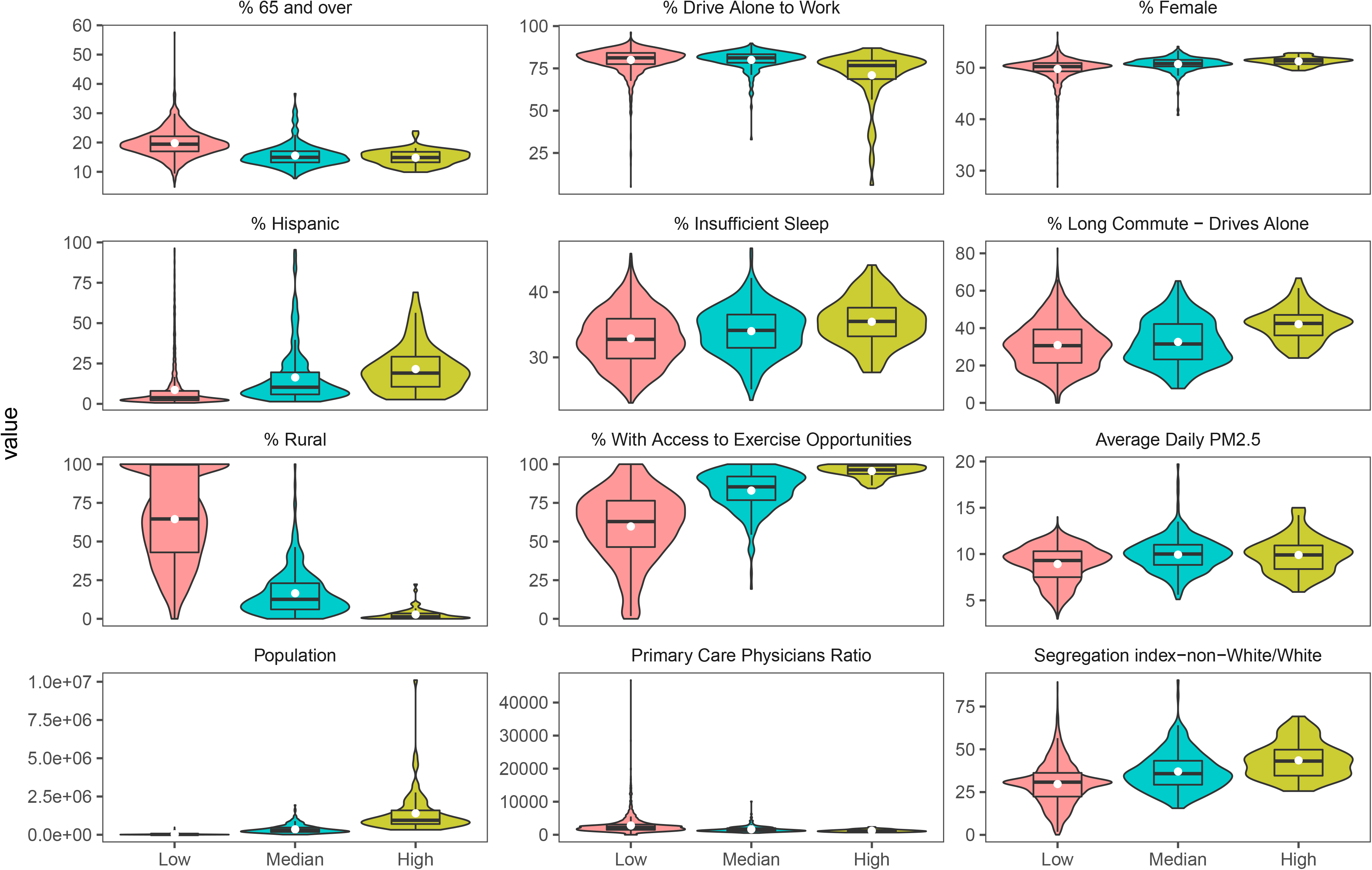
Violin diagram and boxplot of the distributions of the 12 selected risk factors in the three classes of counties. The low, median and high prevalence classes of counties were represented by red, blue and green colors.

### Factors influencing mortality of COVID-19 in the three classes

There were three common factors, namely, residential segregation between non-Whites and Whites, resident population, and the percentage of Hispanic population, which had statistically significant (*P*<0.05) effects on mortality in all classes. The negative binomial model was used to understand the within-class effects of residential segregation between non-Whites and Whites and the percentage of Hispanic population on mortality of COVID-19 as shown in Figure 3. Note that the higher values of both residential segregation between non-Whites and Whites and the percentage of Hispanic population the higher mortality of COVID-19. In the high prevalence class, an increase in both the residential segregation between non-Whites and Whites and the percentage of Hispanic population resulted in more deaths than other two classes of counties.

**Figure 3.**
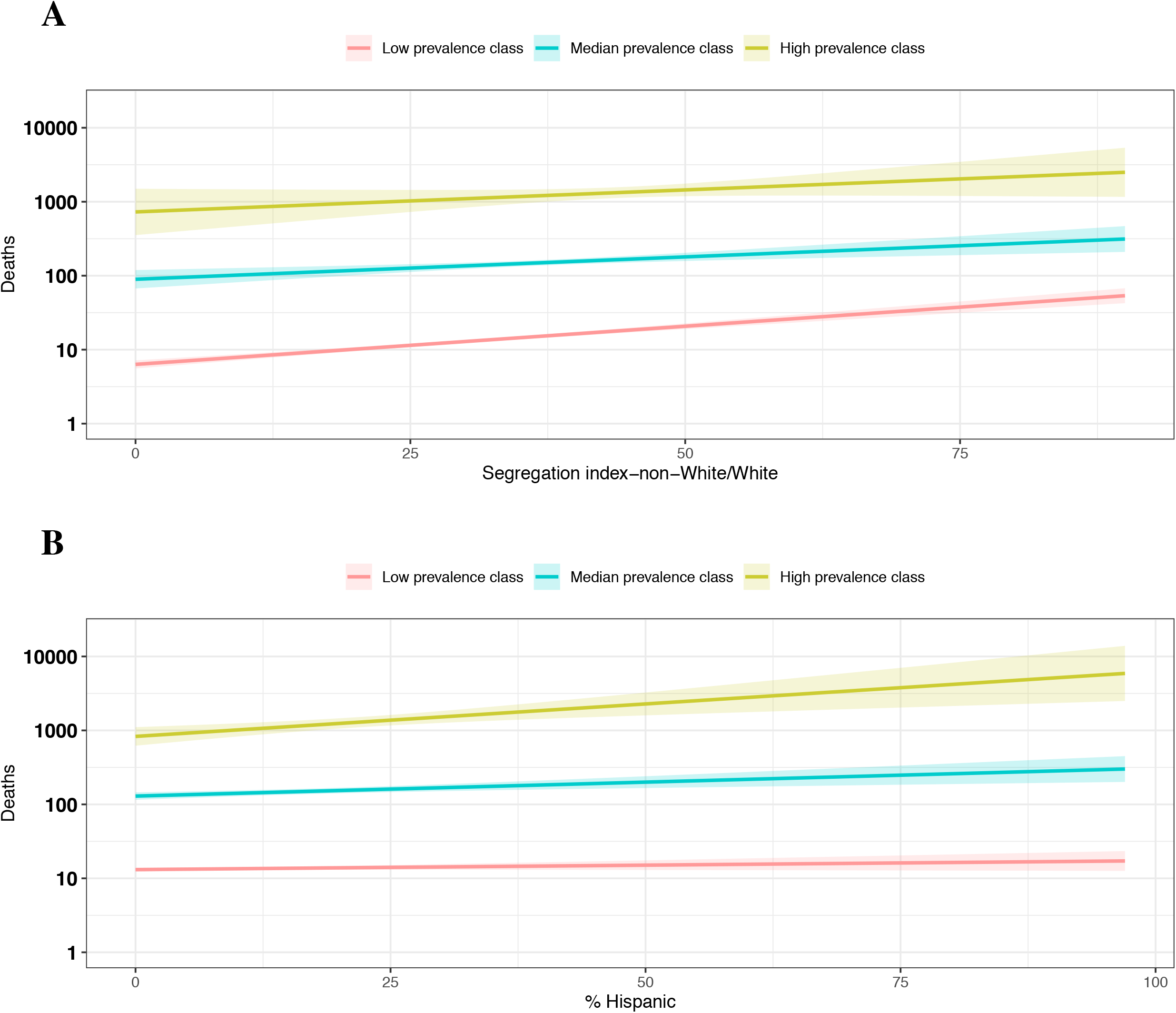
The risk factors common in the three classes of counties. A: Segregation index for non-Whites versus Whites; B: The percentage of Hispanic population. Direct curves were generated using the negative binomial model with the segregation index between non-Whites and Whites or the percentage of Hispanic population as the single covariate.

Table 2 presented the significant factors specific to each class based on the training data. Specifically, in the low prevalence class, nine variables were significantly associated with the mortality of COVID-19. Higher values in the average daily density of PM_2.5_ (0.153, *P*<0.001), the percentage of workforce driving alone to work (0.039, *P*<0.001), the percentage of workforce that had more than 30 minutes commute driving alone (0.015, *P*<0.001), the percentage of adults who reported less than average 7 hours sleeping (0.073, *P*<0.001), resident population (*P*<0.001), the percentage of Hispanic (0.020, *P* <0.001) and female population (0.054, *P<*0.001) and segregation index (0.015, *P*<0.001), significantly increased the number of deaths, while more people living in rural areas (−0.014, *P*<0.001) decreased the number of deaths of COVID-19.

**Table 2.**
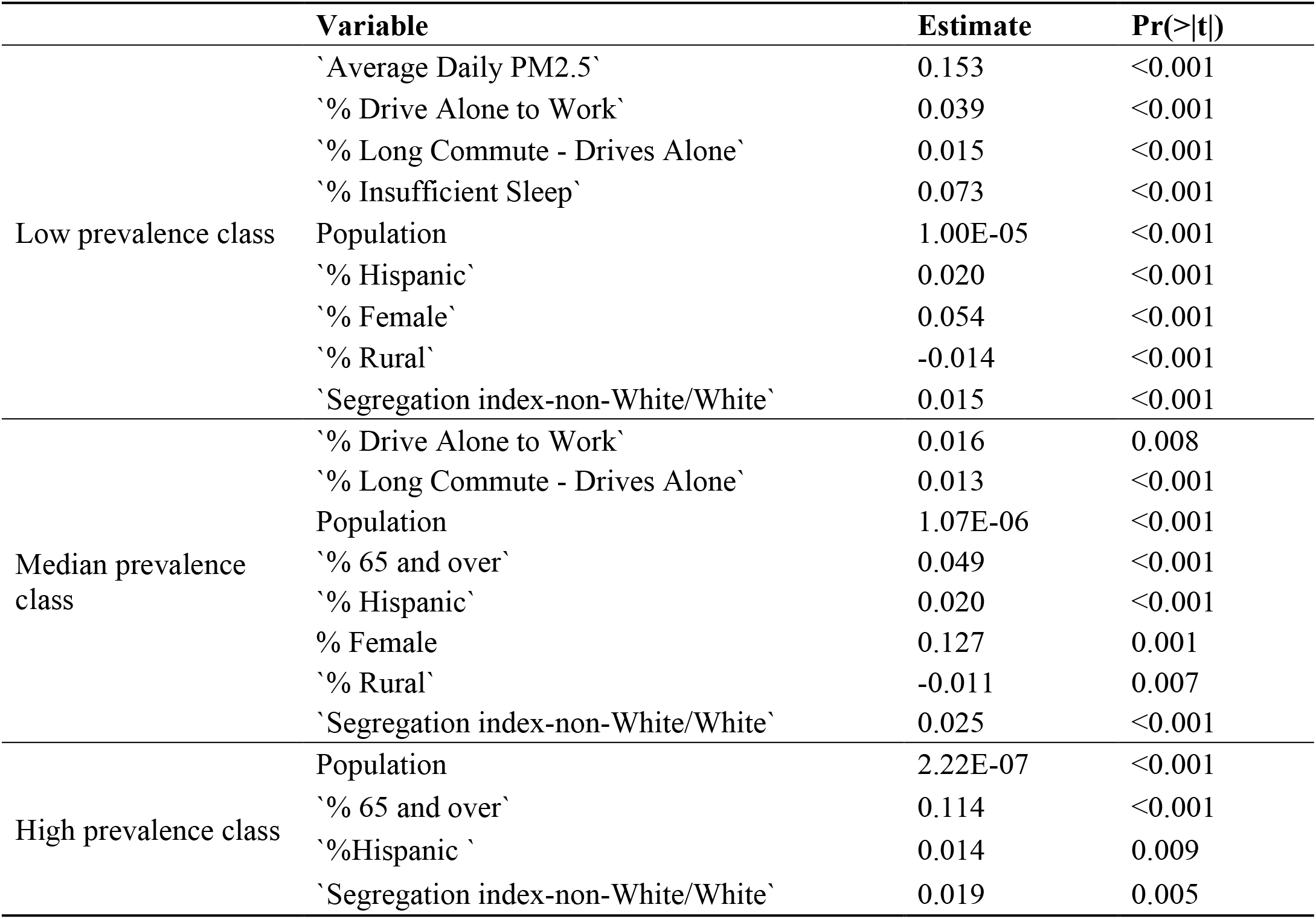
Variables significantly related to the mortality rate of COVID-19 in the three classes of counties.

|  | **Variable** | **Estimate** | **Pr(>\|t\|)** |
| --- | --- | --- | --- |
| Low prevalence class | `Average Daily PM2.5` | 0.153 | <0.001 |
|  | `% Drive Alone to Work` | 0.039 | <0.001 |
|  | `% Long Commute - Drives Alone` | 0.015 | <0.001 |
|  | `% Insufficient Sleep` | 0.073 | <0.001 |
|  | Population | 1.00E-05 | <0.001 |
|  | `% Hispanic` | 0.020 | <0.001 |
|  | `% Female` | 0.054 | <0.001 |
|  | `% Rural` | -0.014 | <0.001 |
|  | `Segregation index-non-White/White` | 0.015 | <0.001 |
| Median prevalence class | `% Drive Alone to Work` | 0.016 | 0.008 |
|  | `% Long Commute - Drives Alone` | 0.013 | <0.001 |
|  | Population | 1.07E-06 | <0.001 |
|  | `% 65 and over` | 0.049 | <0.001 |
|  | `% Hispanic` | 0.020 | <0.001 |
|  | % Female | 0.127 | 0.001 |
|  | `% Rural` | -0.011 | 0.007 |
|  | `Segregation index-non-White/White` | 0.025 | <0.001 |
| High prevalence class | Population | 2.22E-07 | <0.001 |
|  | `% 65 and over` | 0.114 | <0.001 |
|  | `%Hispanic` | 0.014 | 0.009 |
|  | `Segregation index-non-White/White` | 0.019 | 0.005 |

In the median prevalence class, eight variables were significantly associated with the deaths of COVID-19. Higher values in the percentage of workforce driving alone to work (0.016, *P*=0.008), the percentage of workforce that had more than 30 minutes commute driving alone (0.013, *P*<0.001), resident population (*P*<0.001), the percentage of population aged over 65 (0.049, *P*<0.001), the percentage of Hispanic (0.020, *P*<0.001) and female population (0.127, *P*=0.001) and segregation index (0.025, *P*<0.001)-led to an increase in deaths as opposite to a decrease in deaths of COVD-19 for more people living in rural areas (−0.011, *P*=0.007) .

In the high prevalence class, four variables were significantly associated with mortality. Higher values in resident population (*P*<0.001), the percentage of population aged over 65 (0.114, *P*<0.001), the percentage of Hispanic population (0.014, *P=*0.009) and segregation index (0.019, *P*=0.005), caused more deaths.

For each class of counties, the model obtained from the training data was employed to predict the deaths of COVID-19 on August 27, 2020 using the testing data. The corresponding RMSE values for the mortality ratio were 0.056%, 0.041%, and 0.088%, respectively, in the low, median, and high prevalence classes.

## Discussion

Using the time trends of the cumulative confirmed cases in 3,125 counties in the United States, we categorized those counties into three levels of infection. The low prevalence class counted for 88% of the 3,125 counties. Their resident population was remarkably smaller than the other two classes of counties. But the resident population size increased the mortality of COVID-19 regardless of the level of COVID-19 prevalence. A higher population density may increase more contacts in social distancing[17, 18], leading to a higher risk in mortality of COVID-19. On the contrary, a higher percentage of residents living in rural areas in both the low and median prevalence classes of counties may reduce the mortality. Disparities in race and ethnicity were found in the infected populations. For example, Blacks were reported to be prone to COVID-19[19, 20], and living settings of racial/ethnic minorities were founded to be more crowded, making social distancing difficulty [21]. In this study, we found that Hispanics were more vulnerable. Further investigation is warranted to study the racial disparity in the mortality of COVID-19. However, the segregation index between non-Whites and Whites revealed the racial disparity in health, leading to differences in health status not only at the individual level but also at the community level[22]. A higher values in the segregation index indicated the poor health status, which may increase the mortality of COVID-19[17]. This health inequality increased the mortality rates of COVID-19 in all classes of counties.

For the low and the median prevalence class of counties, more workforce driving alone to work and commuting long-distance may increase the levels of anxiety[23], leading to the high mortality in COVID-19. A higher percentage of long-distance commuting workforce was also linked to a high level of anxiety for commuters[23]. And substantial time spent by long-distance commuters could inhibit their healthy behaviors[24]. The stress and less healthy behaviors may increase individual’s vulnerability to COVID-19[27-29]. Also, long-distance commute may be necessary for people who work in relatively higher dense areas where the risk of COVID-19 is high.

The counties in both the low and the median class of prevalence were accounted for 97.44% of the infected counties, where the higher values in the percentage of female population increased the mortality of COVID-19.

The percentage of adults with inadequate sleeping time was found to increase the mortality of COVID-19 in the low prevalence class of counties. Sleeping time was reported to be associated with the health system[25]. The higher number of people who had inadequate sleeping time, the more adverse effects of sleep on immunity were identified[26]. The air quality also was reported to be associated with the mortality rate of COVID-19[10, 30, 31].

For both the median and the high prevalence class of counties, there was an age trend in the mortality rate of COVID-19. In those counties, there was a higher percentage of elderly, indicating a larger population of individuals aged over 65, which increased the mortality rate of COVID-19[11].

## Conclusion

This study identified several significant risk factors associated with the mortality of COVID-19, and our findings are highly valuable and timely for the decision-makers to develop strategies in reducing the mortality of COVID-19. The study relied on mortality data on August 27, 2020. The counties were randomly divided into the training and testing data once. However, we offered the epidemiological picture to facilitate the identification of important factors influencing the mortality of COVID-19 across different levels of infected counties in the United States. Regardless of the regions, the factors linked to the poor health status contributed to higher mortality of COVID-19. Improving the clinical care and eliminating the racial health inequality, combined with improving physical environment were expected to significantly decrease the mortality rate of COVID-19. Thus, we recommended that local governments should reduce physical and psychological risks in residential environments.

## Data Availability

We collected the number of cumulative confirmed cases and deaths from March 1 to August 27, 2020, for counties in the United States from the New York Times. The COVID-19 confirmed cases and deaths were identified by the laboratory RNA test and specific criteria for symptoms and exposures from health departments and U.S. Centers for Disease Control and Prevention (CDC). The county health rankings reports from year 2020 were compiled from the County Health Rankings and Roadmaps program official website. There were 77 measures in each of 3142 counties, including the health outcome, health behaviors, clinical care, social and economic factors, physical environment, and demographics. We refer to the official website of the County Health Rankings and Roadmaps program for detailed information.

https://www.nytimes.com/interactive/2020/us/coronavirus-us-cases.html.

https://www.countyhealthrankings.org/reports

https://www.cdc.gov/coronavirus/2019-ncov/cases-updates/cases-in-us.html

## Declarations

### Ethics approval and consent to participate

Not applicable

### Consent for publication

Not applicable

### Availability of data and materials

The data that support the findings of this study are available from the New York Times and the County Health Rankings and Roadmaps program website. The data and R files supporting the conclusions of this article are available in the https://github.com/tingT0929/Risk-factors-associated-with-mortality-of-COVID-19.

### Authors’ contributions

TT, XW and HZ developed the idea and research. TT, JZ, LH and YJ wrote the first draft of the manuscript and all other authors discussed results and edited the manuscript. JZ, LH, and YJ collected and validated epidemiological data. CD and ZL collected and compiled census data. All authors read and approved the final manuscript

## Acknowledgements

We would like to thank all individuals who are collecting epidemiological data of the COVID-19 outbreak around the world.

